# Long-Term Healthcare Utilization After Genomic Diagnosis in Seriously Ill Children

**DOI:** 10.64898/2026.02.24.26345973

**Authors:** Joao M. L. Dias, Ravi P. More, Duncan Butler, Clare Aldus, Julian Brown, Courtney E. French, Helen Dolling, F. Lucy Raymond, David H Rowitch, Catherine E Aiken

## Abstract

**Importance:** Whole genome sequencing (WGS) is increasingly used to diagnose severely ill children, yet the long-term impact of a genetic diagnosis on healthcare utilization and resource allocation remains poorly understood.

**Objective:** To determine the influence of a genetic diagnosis via WGS on long-term healthcare utilization metrics in severely ill children.

**Design:** A retrospective cohort study using data from the Next Generation Children study (2016-2020) with record linkage and analysis of primary care records conducted between 2022 and 2024.

**Setting:** A multicenter study involving primary care and hospital records linked via the UK

National Health Research Institute (NIHR) Rare Disease Bioresource, Cambridge, UK.

**Participants:** A referred sample of 270 severely ill children who underwent WGS.

**Exposure(s):** Receipt of a genetic diagnosis (87/270; 32%) compared to those who remained undiagnosed (183/270; 68%) following WGS.

**Main Outcome(s) and Measure(s):** Comparison of 36 healthcare utilization parameters, including hospitalizations, primary care prescriptions, and diagnostic tests.

**Results:** Among the 270 children analyzed, those receiving a genetic diagnosis (n=87) exhibited significantly higher overall healthcare utilization compared to undiagnosed peers (n=183). This included increased hospital admissions and outpatient visits, particularly for neurodevelopmental and seizure-related conditions. Diagnosed children received a higher volume of neurological, gastrointestinal, and nutritional prescriptions. The most pronounced differences in utilization were observed in children initially diagnosed in neonatal (NICU) or pediatric (PICU) intensive care settings. While genetic diagnosis was not associated with reduced healthcare costs during the study period, it was linked to more targeted, condition-specific medical care.

**Conclusions and Relevance:** WGS diagnosis facilitates the integration of specialist care and the alignment of healthcare resources with the specific needs of children with complex disorders. These findings suggest that while costs may not decrease immediately, a diagnosis enables more precise and targeted clinical management.

**Key Points:** *Question:* Does a genetic diagnosis through whole genome sequencing influence long-term healthcare utilization in severely ill children?

*Findings:* In this cohort study of 270 children, those who received a genetic diagnosis demonstrated significantly greater overall healthcare utilization, including more hospitalizations and targeted prescriptions, compared with undiagnosed children.

*Meaning:* A genetic diagnosis facilitates the integration of specialized, condition-specific care, helping to align healthcare resources with the individual needs of children with complex disorders.

## Introduction

Children admitted to Neonatal or Paediatric Intensive Care Units (NICU/PICU) often present with severe, life-threatening conditions affecting short-term survival and long-term neurodevelopment ^1–3^. Genetic disorders are now recognised as leading contributors to morbidity and mortality, affecting 10-30% of children admitted to NICU/PICU ^4–11^. This supports the use of whole genome sequencing (WGS) in clinical practice, particularly for cases of suspected rare or monogenic disease ^6,12–14^.

Prior health economic analyses have indicated that WGS is cost-saving in the PICU setting, largely due to reductions in the length of the acute hospital stay and the avoidance of invasive procedures ^2–4,8,15–18^. Initiatives such as Project Baby Bear in California report short-term net healthcare savings exceeding $14,000 per ICU infant tested ^19^. In contrast to studies of short-term financial benefits, the long-term implications of genomic diagnosis for healthcare utilization remain under-explored, particularly in the UK national health service (NHS) ^20,21^. We address this knowledge gap by linking paediatric WGS results to clinical outcome data provided by NHS Prescribing Services (ECLIPSE Live), which comprises linked primary and secondary care data for >25 million NHS patients ^22^.

The Next Generation Children’s (NGC) project, conducted in Cambridge (2016-2020), was an early demonstration of utility of WGS in the intensive care setting, with more than 90% of clinicians reporting improved confidence in patient management and family communication following testing ^23^. In the NGC, 25-46% of critically ill children (NICU/PICU) received a molecular diagnosis from WGS ^12^. The NHS has subsequently provided rapid genome sequencing (R14 service) for acutely ill children who meet testing criteria ^24,25^, impacting patient management and/or family reproductive counselling in nearly all diagnosed cases^20,26^.

This study incorporated detailed clinical phenotypes and NHS primary care records (ECLIPSE Live) from electronic records alongside genomic findings from NGC project whose families had consented for linkage research as a part of the UK NIHR Rare Disease Bioresource. We examined whether long-term healthcare use patterns differed between children who received a diagnosis via WGS compared to those who did not. Secondly, we evaluated the possibility that healthcare utilization and other clinial characteristics might be an independent means to identify children with a higher probability of a positive genetic result from WGS.

## Methods

### Cohort Description

Participants (n=270) were part of the NGC project; a cohort of seriously ill children recruited from the NHS between December 2016 and August 2020 ^23^. NGC was designed with broad inclusion criteria for recruitment from NICU/PICU, paediatric neurology and genetics clinics. Peripheral blood samples were collected from probands and, where possible, both biological parents for trio analysis. Subjects included babies and children with congenital anomalies, neurological symptoms (including seizures), suspected metabolic disease, extreme intrauterine growth restriction, and unexplained critical illness. The NGC project is described in detail elsewhere ^12,23^.

### Demographics and Clinical Phenotype Data

Clinical phenotype data from the electronic medical records were obtained via the NIHR BioResource for Translational Research in Common and Rare Diseases. This resource included standardised demographic information, Human Phenotype Ontology (HPO) terms, family pedigrees, and detailed medical history ^12,23^. For this analysis, HPO terms deeper than level three in the ontology hierarchy were mapped to their corresponding system-level parent terms, yielding a consolidated set of 23 unique HPO categories. This transformation was performed using the ontologyIndex R package and the latest hp.obo release (hp/releases/2025-05-06), with obsolete terms updated via the Monarch ^27^.

### DNA Variant Classification

DNA variants were classified pathogenic (P), likely pathogenic (LP), variants of uncertain significance (VUS) or genetically undiagnosed as previously described ^23^.

### Primary Care Records

Primary care records were accessed through the ECLIPSE Live platform (Prescribing Services Ltd) ^22^. Due to the absence of complete long-term data, cases were excluded if the child deceased (except for one instance where re-consent was obtained), withdrew consent, had missing identifiers, or changed General Practitioner (GP) prior to the linkage date (see Figure 2A for full cohort diagram). Primary care data in the UK are controlled by individual practices, which provided permission to access data for consenting participants. Identifiers, including NHS number, date of birth, and sex were used to securely link the datasets. Linked data were pseudonymised and accessed via a secure data environment (SDE). The available dataset included healthcare usage and associated costs collected from January 2022 to June 2024. The data were sorted into four categories: outpatient and Emergency Department visits, prescriptions, pathology reports, and clinical conditions, and processed into 36 primary care features for further analysis (Table 1). HPO terms (n=657) for the analytic cohort were also identified, with the 3 most common selected for further analysis: hypotonia, seizures, and developmental delay. A comprehensive feature dictionary and derivation methodology are included in the Supplementary Information.

**Table 1.** Primary care features used in the analysis, including outpatient visits, prescriptions, pathology, and conditions. The table also lists the selected conditions identified from the HPO terms.

| Category | Details | Category | Details |
| --- | --- | --- | --- |
| **Outpatients visits and A&E admissions (6)** | • Number of Outpatients visits<br>• Number of A&E visits<br>• Number of admissions<br>• Number of admission days<br>• Number of unique specialties<br>• Total cost of visits and admissions | **Prescriptions (22)** | • Number of unique prescriptions types<br>• Number of prescriptions<br>• Total cost treatment<br>• Number of drug groups<br>• allergology<br>• algology<br>• cardiology<br>• care/hygiene<br>• dental<br>• dermatology<br>• diagnostic<br>• endocrinology<br>• gastroenterology<br>• immunology<br>• lines, tubes, and catheters<br>• hepatology<br>• neurology<br>• nutrition<br>• pneumology<br>• short-term Infection<br>• steroid<br>• urology and gynaecology<br>• vaccination |
| **Pathology (3)** | • Number of unique blood test parameters<br>• Number of unique blood tests<br>• Number of unique blood parameter types (nephrology, hematology, hepatology, nutrition, infection or endocrinology) |  |  |
| **Conditions (7)** | • Number of unique conditions<br>• **cardiorespiratory**<br>• **endocrinology and immunology**<br>• **neurodevelopmental**<br>• **sensory**<br>• others<br>• no records |  |  |

### Statistical analysis

Statistical analyses were performed in R (version 4.5.0). Healthcare utilisation variables were compared between genetically diagnosed and undiagnosed groups across whole cohort and predefined clinical conditions (cardiorespiratory, endocrine/immunological, neurodevelopmental and sensory) and phenotypic subsets (hypotonia, seizures and developmental delay). For each comparison the Wilcoxon rank-sum test was applied using the ‘wilcox.test’ function. A stringent threshold was adopted where only comparisons with a p-value below 0.001 and at least 7 observations per group were considered significant. This strict criterion was chosen to increase confidence in the observed differences and reduce the likelihood of false positives. Median values for each group were calculated using the ‘median’ function with missing values removed. Table S1 shows the distribution of cases across the full cohort and within each clinical setting, for each diagnostic group and for each condition highlighted in bold in Table 1.

Results were visualized using heatmaps generated with the ‘Heatmap’ function from the ‘ComplexHeatmap’ package. The top and bottom position within each heatmap cell was determined based on the median value in each subset to indicate which group had higher utilization. Violin plots were created using ‘ggplot2’ with the ‘geom_violin’ function combined with ‘facet_wrap’ for grouping. A pseudo log scale (‘pseudo_log’) was applied to the y-axis to allow better visualization of results when healthcare utilization variables differed by orders of magnitude.

### Predictive Model Development

To estimate the likelihood of a genetic diagnosis based on phenotypic data, we developed a predictive model integrating HPO terms, primary care records, and demographic variables. For each participant, a binary matrix of organ-level HPO terms (23 terms) was constructed. Demographic variables included age, sex, ethnicity, Index of Multiple Deprivation (IMD) score, and clinical setting (e.g., NICU or PICU). All features were combined into a single binary matrix for modelling. Numeric features were divided into four quantile-based intervals using breakpoints at the 0th, 25th, 50th, 75th, and 100th percentiles, rounded up for interpretability. Features with four or fewer unique values were one-hot encoded. Missing values were encoded as zeros. Figures 1 and S1 shows the histograms of demographic and primary care features across 270 cases, stratified by diagnostic group, with quantile-based bins for standardized comparisons. Dimensionality reduction was performed using Principal Component Analysis (PCA) via ‘*prcomp*’ (base R), applied without centering or scaling. Linear Discriminant Analysis (LDA) was then fitted on the PCA components using ‘*lda*’ from the ‘*MASS*’ package (v7.3.65). Model stability and performance were assessed using bootstrap resampling with repeated random splits (70% training, 30% testing) via ‘*sample.split*’ from ‘*caTools*’ (v1.18.2). Models were trained on up to 70 principal components. Performance metrics included sensitivity, specificity, and ROC curves computed with ‘*roc*’ from ‘*pROC*’ (v1.18.5). AUC values were aggregated across bootstrap iterations. The final model was selected based on the highest mean AUC in the validation dataset.

**Figure 1.**
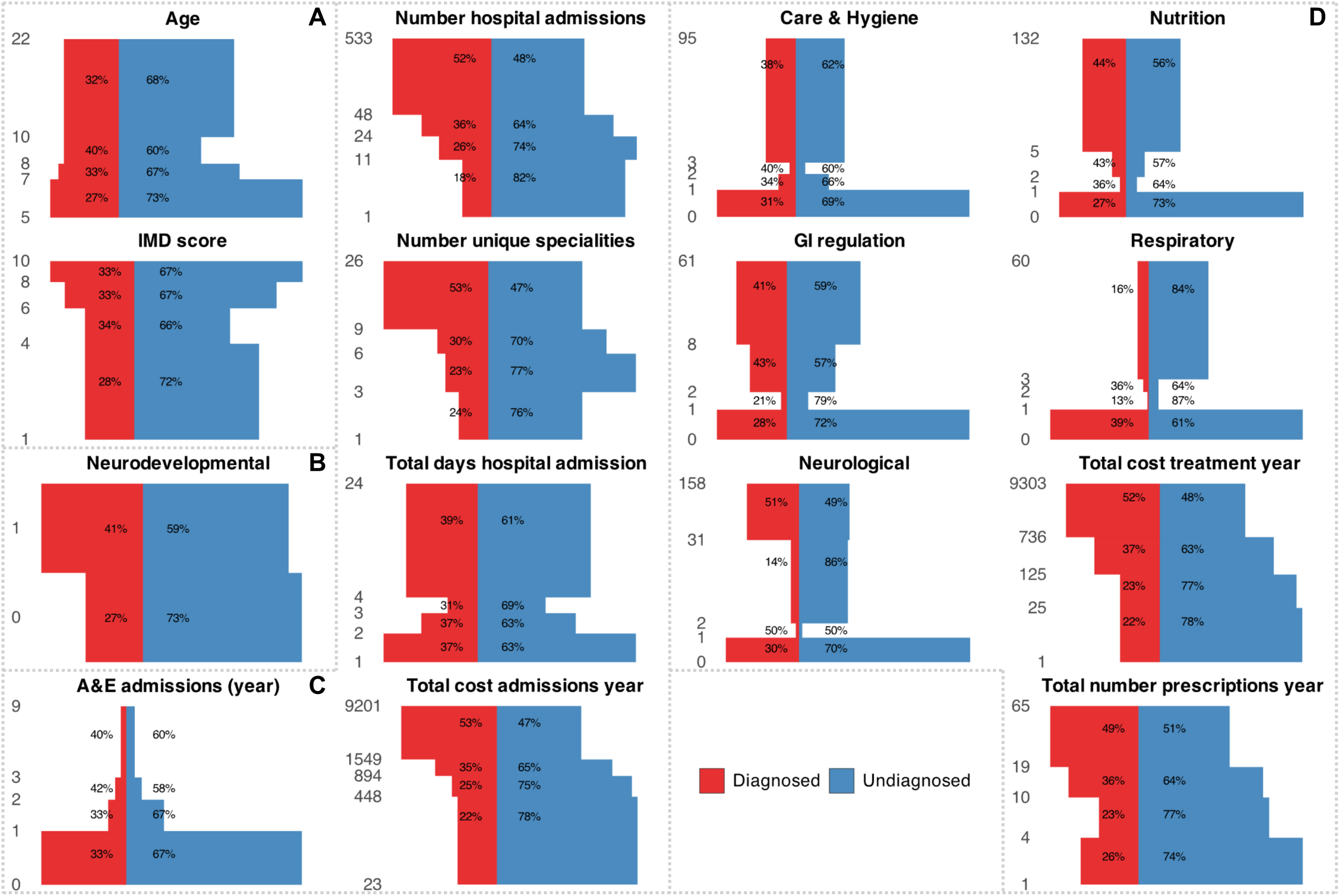
Histogram of statistically significant demographic and primary care features across 270 cases, stratified by diagnostic group. Each feature is discretized into four quantile-based bins to enable standardized comparisons across variables. Bars indicate the absolute number of individuals per bin, grouped by diagnostic status (diagnosed vs. undiagnosed). To enhance visibility for features with high or skewed values, the y-axis is scaled using log1p.

**Figure 2:**
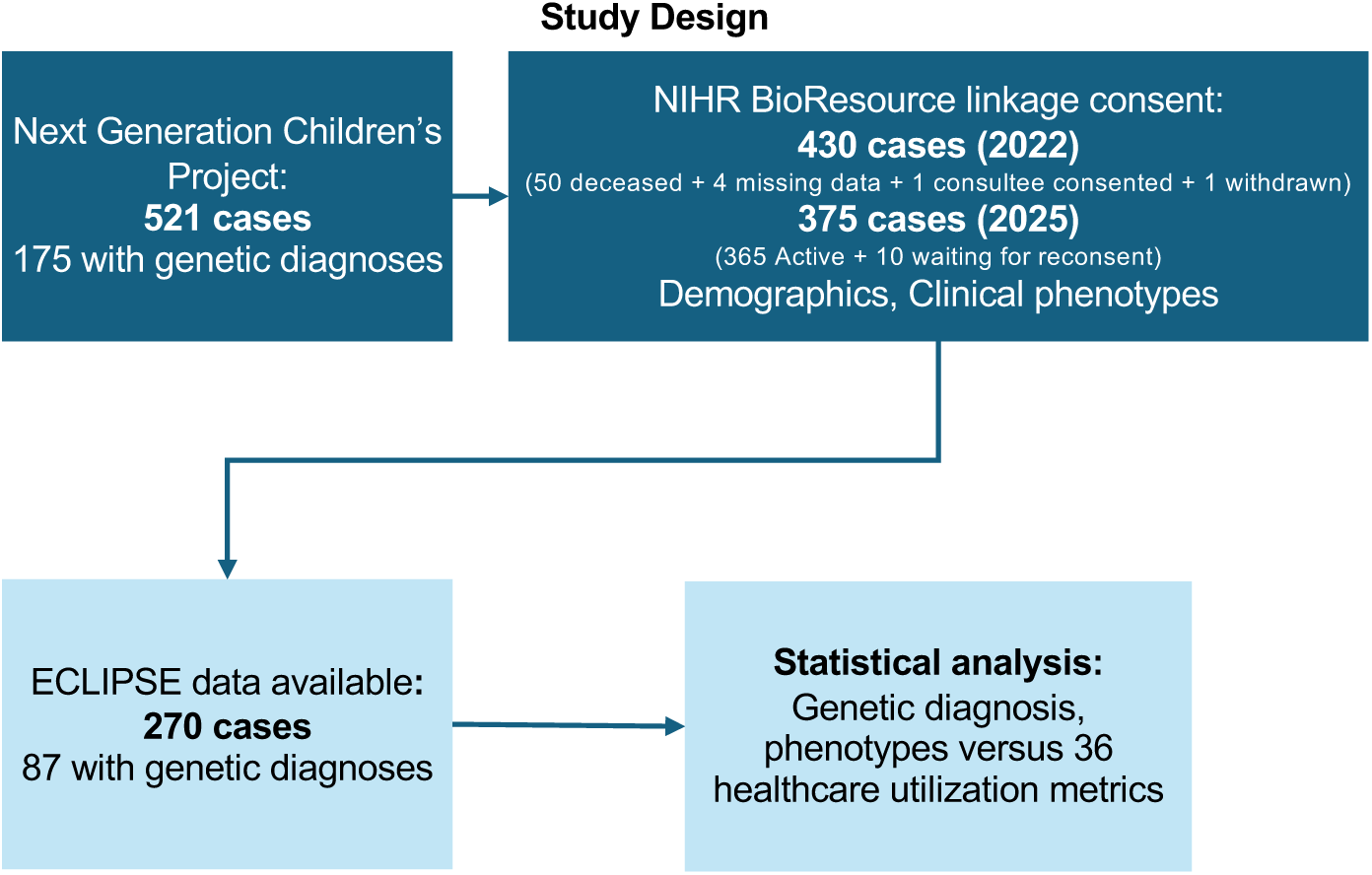
Flowchart of participant selection and data linkage. Overview of the study cohort derivation from the Next Generation Children’s (NGC) project (n=521). Participants were sequentially filtered based on consent for data linkage via the NIHR BioResource and the availability of linked longitudinal primary care records in the ECLIPSE database. The final analytical cohort comprised 270 children (87 with a molecular diagnosis and 183 undiagnosed), who were assessed against 36 healthcare utilisation metrics.

### Feature Impact Analysis

To assess the contribution of individual features, we applied a leave-one-feature-out approach using the final PCA+LDA models. For each feature, its contribution to the PCA projection was removed by subtracting its loading from the original components. Updated scores were passed through the trained LDA model to recalculate probabilities. The difference between original and modified probabilities was summarized as mean probability change with 95% confidence intervals (±1.96 × SE). Features with absolute mean differences >0.02 were considered influential. Results were visualized as volcano plots, with the x-axis representing mean probability difference and the y-axis the confidence interval width. Features were color-coded by effect direction and labelled when significant.

Plots were generated using ‘*ggplot2*’ (v3.5.2) and ‘*ggrepel*’ (v0.9.6). All analyses were performed in R (v4.3.2), with full implementation details in the Supplementary Methods. Results were visualized using heatmaps generated with the ‘*Heatmap*’ function from the ‘*ComplexHeatmap*’ package (v2.24.1). The top and bottom position within each heatmap cell was determined based on the median value in each subset to indicate which group had higher utilization. Violin plots were created using ‘*ggplot2*’ (v4.0.1) with the ‘*geom_violin*’ function combined with ‘*facet_wrap*’ for grouping. A pseudo log scale (‘*pseudo_log*’) was applied to the y-axis to allow better visualization of results when healthcare utilization variables differed by orders of magnitude.

### Research Governance

Ethical approval for the NGC study was granted by the Cambridge South Research Ethics Committee (13/EE/0325). Written informed consent was obtained from parents or legal guardians for diagnostic whole-genome sequencing (WGS) and data linkage to health records via the NIHR BioResource for Translational Research in Common and Rare Diseases (RG94028). The specific linkage to primary care data was approved by the NIHR Bioresource Data Access Committee (DAA179).

## Results

### Cohort Characteristics and Clinical Context

The NGC study enrolled 521 probands and families (trios), of whom 375 consented to data linkage. Among these, 270 cases (72%) could be successfully linked to ECLIPSE Live data, forming the analytical cohort (Figure 2A). All cases (n=270) underwent WGS, with 87 (32%) receiving a genetic diagnosis. Table S2 shows the distribution of age at recruitment, gender, ethnicity, IMD score, clinical setting, and genetic diagnosis. Forty-five percent were female. Most of the participants were European (83%), with 7% had South Asian background; individuals from African and Finnish-European backgrounds made up less than 1%, and 9% came from other backgrounds. Thirty percent of the cohort were in IMD deciles 1 to 4 (lowest socio-economic status group). Children were recruited from clinical settings that included neonatal units (35%), neurodevelopmental units (41%), and paediatric critical care (24%). The genetic diagnostic yield in the analytic cohort was 32%, similar to the full NGC cohort (34%), with 68% undiagnosed. The analytic cohort (recruited between 2016-2020) consisted of older children (analysed in 2025) with a slight male predominance and higher deprivation compared to the NGC overall (Supplemental Table 2).

### Healthcare Utilization and Cost Patterns

Diagnosed cases consistently had higher healthcare utilization (p<0.001 across comparisons; Figure 3). Figures S2–S5 provide detailed comparisons of all statistically significant differences across conditions and utilization metrics. On group level, diagnosed cases experienced more hospital admissions (median 37 versus 22), more unique specialties (8 versus 5), more primary and secondary care outpatient visits each year (14 versus 8), and higher annual admission costs (£1,281 versus £795). These trends were particularly prominent in children with hypotonia, seizures, and developmental delay (Figure S2C-E), although seizure cases did not show a difference in the number of specialties. Annual treatment costs were also higher for diagnosed cases (£335 versus £77), especially for genetic neurodevelopmental (£1,280 versus £130) (Figure S2G) and seizure-related conditions (£1,277 versus £60) (Figure S2I). Diagnosed seizure cases had more prescriptions, particularly for neurological and gastrointestinal care. Developmental delay cases showed an increase in prescriptions for hygiene, nutrition, and care support, for example catheters, feeding tubes, positioning aids (Figure S2J).

**Figure 3:**
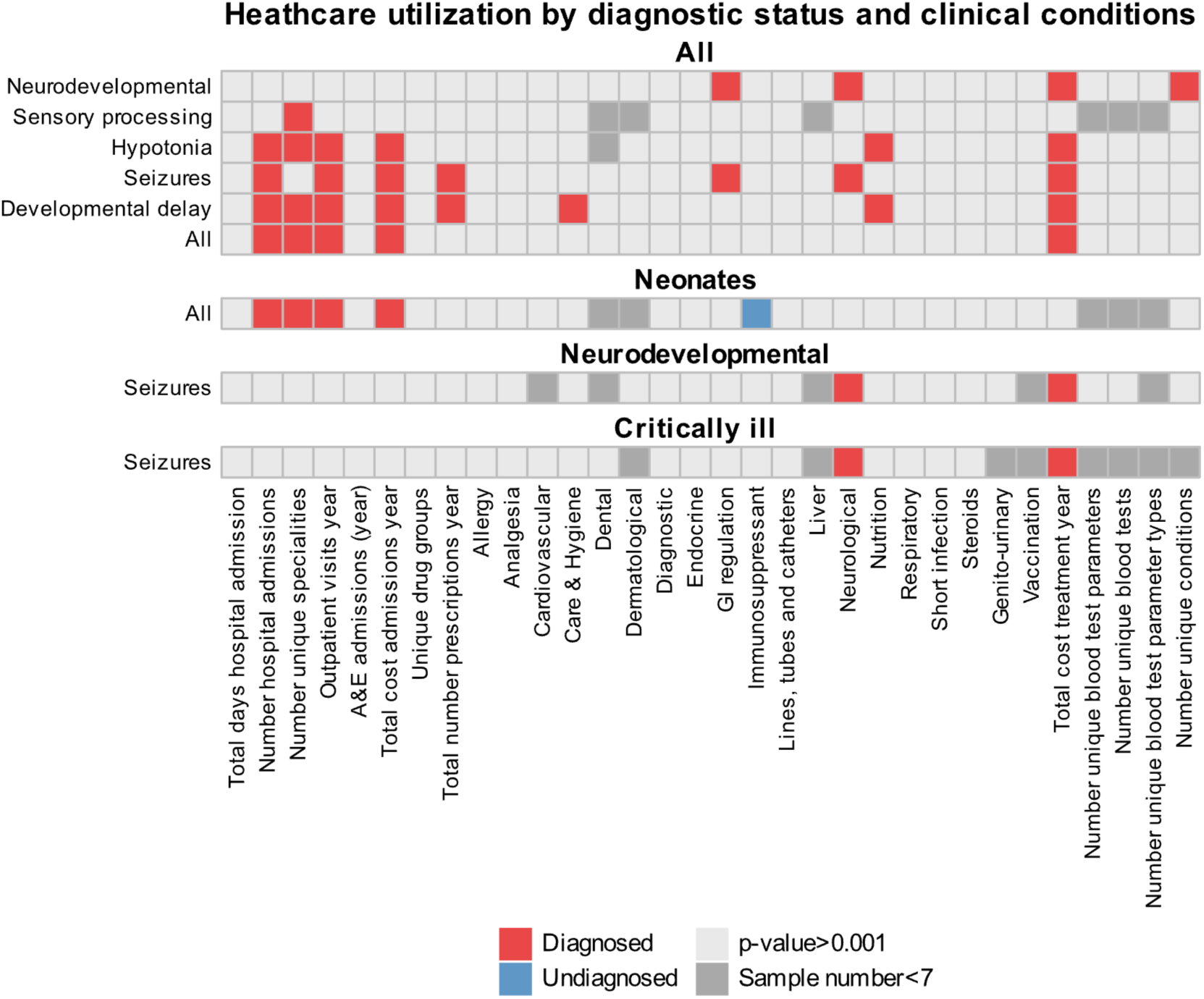
Increased healthcare costs and resource utilization in diagnosed cases. Heatmaps highlighting higher healthcare utilization across multiple clinical conditions, stratified by diagnosis status, for the full cohort and within the Neonates, Neurodevelopment, and Critically Ill subsets. For each condition and utilization metric, Wilcoxon rank-sum tests were performed to compare diagnosed and undiagnosed cases. The heatmaps use cells to indicate the group (diagnosed or undiagnosed) with the higher mean. Cells with non-significant p-values (p > threshold) are shaded light grey, while those with insufficient sample sizes are shaded dark grey.

Figure 4 shows the healthcare costs associated with a diagnosis, highlighting the higher total admission and treatment costs for diagnosed cases, especially in seizure cases where the cost gap was most noticeable. In children recruited from neonatal units, diagnosed cases had significantly more re-admissions (58 versus 16), specialty involvement (12 versus 5), outpatient visits (21 versus 6), and higher admission costs (£2,092 versus £716) (Figure S3). In neurodevelopmental and PICU settings, the differences focused mainly on neurological prescriptions (42 versus 0) and treatment costs (£1,045 versus £38) (Figure S4). In critical care seizure cases, neurological prescriptions (82 versus 0) and treatment costs (£2,438 versus £72) were significantly higher (Figure S5).

**Figure 4:**
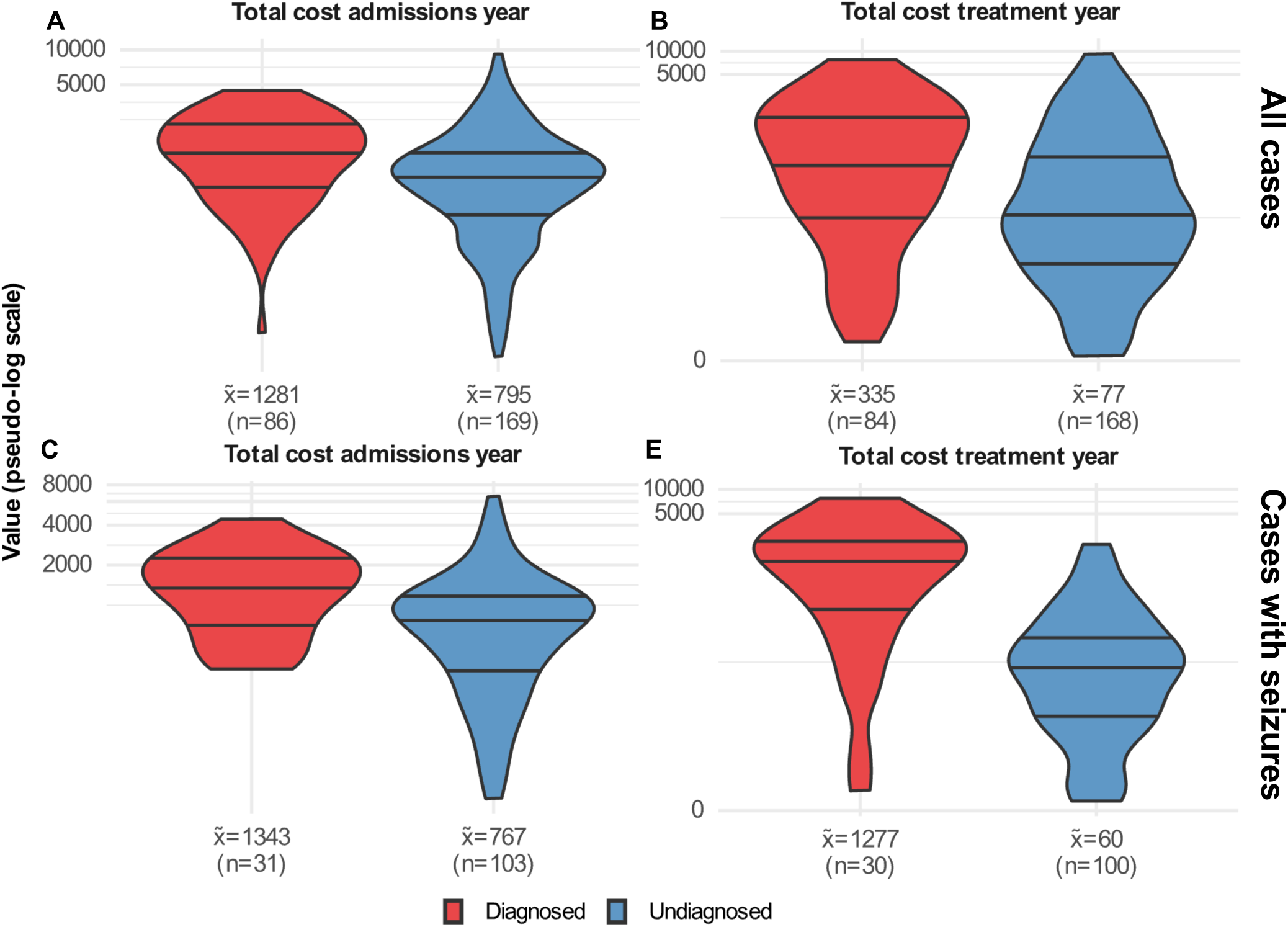
Increased healthcare costs and resource utilization in diagnosed cases. A) Violin plots highlights higher total cost of admission per year for the full cohort (A) and cases with seizures (C), total cost of treatment per year for the full cohort (B) and cases with seizures (E), and number of prescriptions for neurological conditions (D). Each panel corresponds to a specific condition-utilization comparison, with violin plots contrasting diagnosed (red) and undiagnosed (blue) groups. The 0.25, 0.5, and 0.75 quantiles are marked in the plot. The y-axis uses a pseudo-log scale to accommodate wide-ranging values. Median and sample size for each group are annotated below the respective plots.

These findings indicate that diagnosed cases consistently exhibited significantly higher healthcare utilization and costs across all metrics, including hospital admissions, outpatient visits, and prescription costs, with the most pronounced differences observed in children with seizures, hypotonia, and developmental delay.

In order to ascertain whether healthcare utilization patterns could inform the likelihood of obtaining a diagnosis via WGS, a model was trained on 70% of the cohort and validated on 30%, with eight principal components selected (Figure 5A). LDA scores showed clear separation between diagnosed and undiagnosed cases (p<0.0001; Figure 5B), and ROC analysis (Figure 5C) resulted in an AUC of 0.71 (sensitivity 81%, specificity 60%). Training and full datasets had AUCs of 0.78 and 0.76, respectively.

**Figure 5:**
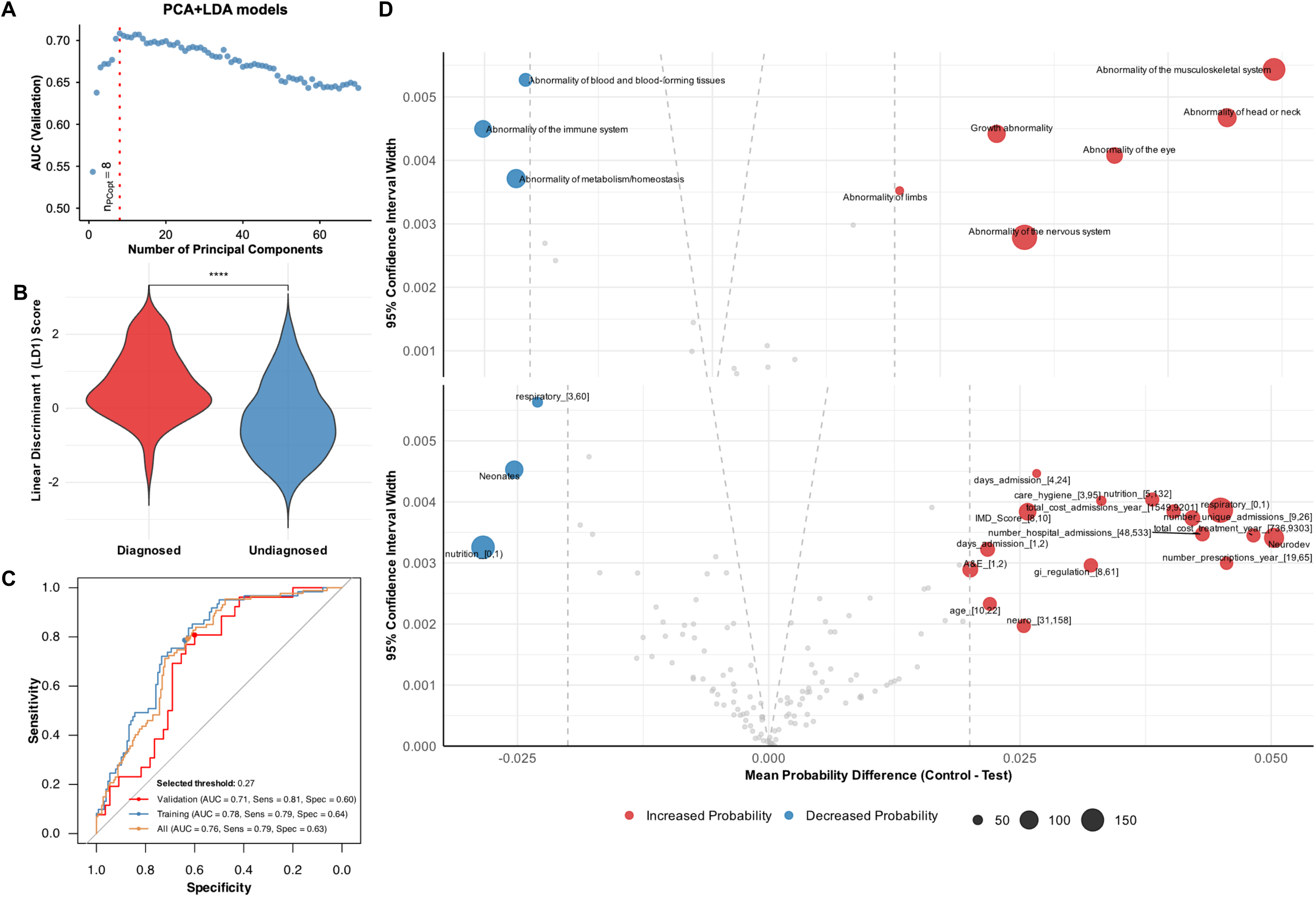
Predictive Modelling of Genetic Diagnosis. A: Average AUC on the validation set as a function of the number of principal components (PCs) for the PCA+LDA models. The red dashed line marks the number of PCs that achieved the highest model performance. B: Violin plots that reveal differences between the diagnostic categories using LDA evaluation of HPO terms. Mann-Whitney U test: p < 0.0001, indicating significant group differences. C: ROC curves for the PCA+LDA model, developed using the optimal number of PCs. The curves for the full (orange), training (blue), and validation (red) datasets are shown. Each curve represents the bootstrap model iteration with an AUC closest to the average AUC from all iterations. The point on each curve indicates the optimal classification threshold that maximizes the sum of sensitivity and specificity. PC: principal Components; AUC: Area Under the Curve; LDA: Linear Discriminant Analysis; ROC: Receiver Operating Characteristic. D: Volcano plot to highlight HPO terms, and demographics and healthcare utilization variable intervals with increased or decreased probability of a genetic diagnosis in the context of a child with cerebral palsy. HPO terms are labelled if the absolute mean probability difference is greater than 0.02 and exceeds the width of its 95% confidence interval. The size of each data point is proportional to each variable frequency in the cohorts. AUC: Area under the curve; HPO: Human Phenotype Ontology; LDA: Linear Discriminant Analysis; VOI: Variant of interest.

Simulation-based feature analysis (Figure 5D) identified key predictors: older age (10 to 22 years), IMD decile 8-10, management in a neurodevelopmental unit, low A&E attendance (1 to 2), high hospital admissions (9 to 26), total admissions (48 to 533), and higher admission costs (£1,549 to £9,201). Both low (1 to 2) and high (4 to 24) inpatient days associated with increased likelihood of diagnosis. Treatment-related predictors included high annual prescriptions (19 to 65), costs (£735 to £9,303), and prescriptions for nutrition (5 to 132), neurology (31 to 158), GI regulation (8 to 61), and hygiene/care (3 to 95). Low respiratory prescriptions (0 to 1) increased likelihood, while high respiratory prescribing (3 to 60), low nutrition prescriptions (0 to 1), and management in a neonatal unit reduced it, likely reflecting incomplete phenotype expression in infancy. Six HPO systems (musculoskeletal, head and neck, growth, eye, limb, and nervous system) were strong predictors, whereas lower likelihood was linked to abnormalities in blood, immune, and metabolic systems. We note that the LDA model, using PCA preprocessing, prioritized clarity but might overlook non-linear effects and increase false positives. In sum, feature analysis of our model revealed that the likelihood of obtaining a positive diagnosis from paediatric WGS in the context of serious illness is strongly associated with high healthcare utilisation, specifically frequent hospital admissions and neurological prescriptions, whereas neonatal presentation and respiratory phenotypes were linked to a lower likelihood of a genetic diagnosis being found.

## Discussion

While several prior studies have indicated benefits and cost-savings of WGS diagnosis for severely ill children in the short term, here we investigated longer term implications of WGS diagnosis for care. We show that combining primary care records and genomic data in seriously ill children provides two main insights. First, cases who received a genetic diagnosis through whole-genome sequencing (WGS) utilised significantly more healthcare resources than undiagnosed peers, especially those with neurodevelopmental and seizure-related conditions. The level of symptoms derived from the HPO terms was equivalent between the diagnosed and undiagnosed groups, however children who received a diagnosis had enhanced usage of specialist pathways and personalised treatment. WGS-diagnosed cases showed increased hospital admissions, outpatient visits, specialist involvement and treatment spending.

Such findings are important for informing clinical services. Our previous work suggests that early detection could help prioritize rapid sequencing and faster management within NHS workflows in the acute illness phase ^12,23^. We now show that diagnosed cases can often access more specialized care after the acute phase, highlighting the importance of recognizing their complex needs in areas like neurology, nutrition, and gastrointestinal support. While this might not lower immediate costs, it optimizes long-term care for chronic neurodevelopmental disorders. We note that none of the patients in our cohort were recipients of gene therapy, such as anti-sense oligonucleotides, which typically require repetitive dosing at high annual cost. Our findings of higher healthcare utilization and cost are thus independent of considerations around advanced therapies, which is an important future consideration.

Healthcare utilization patterns were closely tied to phenotype-diagnosis relationships. Simulation-based feature analysis showed that older age, higher socio-economic status, and some aspects of prescribing and admissions were linked to a higher diagnostic yield. On the other hand, management in neonatal units and respiratory prescribing were associated with a lower yield, possibly due to incomplete phenotype expression or common non-genetic respiratory issues. HPO terms related to musculoskeletal, head and neck, growth, eye, limb, and nervous system problems were the strongest predictors. Diagnosed cases showed higher healthcare utilization and costs, especially in seizure-related and neonatal admissions. More neurological prescriptions in neurodevelopmental and critical care groups reflected focused management of chronic needs following diagnosis.

### Limitations

A key strength of this study is the connection of prospectively captured phenotypes from a WGS program with long-term primary care data, providing a broader view of long-term healthcare needs. We were able to generate a predictive machine learning model merging demographics, healthcare usage, and HPO terms. This model demonstrated good performance (AUC 0.71; sensitivity 81%; specificity 60%), suggesting that routinely collected clinical data can help identify children in whom WGS is more likely to yield a monogenic diagnosis. Such tools might in the future be further developed for clinical decision support.

The limitations of this work include a relatively small analytical dataset (270 cases). We were unable to include participants who did not consent for linkage studies at the time of initial recruitment, who had died in the interim period, and who were not registered with a GP practice with available linked data.

## Conclusion

Our findings indicate that WGS diagnosis, while not reducing overall cost, facilitates integration of specialist care for children with complex disorders, helping align healthcare resources with individual needs. Future work should aim to validate our findings across NHS regions and with larger numbers of patinets. Long-term strategies should focus on predicting care intensity and specialty needs, with cost evaluations that include primary, secondary and social care costs.

## Supporting information

Suplementary_Information_dictionary

## Data Availability

All data produced in the present work are contained in the manuscript

## Acknowledgements

We are grateful for the support of the NIHR BioResource for Translational Research and in helping deliver this research study. This research was supported by funding from the Rosetrees Charitable Trust (to D.H.R.), The Isaac Newton Trust (to D.H.R.), Action Medical Research (GN2788; to C.E.A.), a Wellcome Trust Discovery Award (312471/Z/24/Z; D.H.R., C.E.A.) and NIHR Cambridge Biomedical Research Centre (NIHR203312; D.H.R, C.E.A). Views expressed are those of the authors and not necessarily those of the NIHR or the Department of Health and Social Care.

## Contributors

Conceptualization: DHR, CEA

Data curation: JMLD, RPM, HD, DB

Formal analysis: JMLD, RPM

Funding acquisition: DHR, CEA

Investigation: JMLD, RPM, DB, CEA

Methodology: JMLD, RPM, CEA

Project administration: DHR, CEA

Software: JMLD, RPM

Supervision: DHR, CEA

Visualization: JMLD, RPM, DHR

Writing – original draft: JMLD, RPM

Writing – review & editing: JMLD, RPM, CEF, HD, FLR, DB, JB, DHR, CEA

---

**Figure S1.**
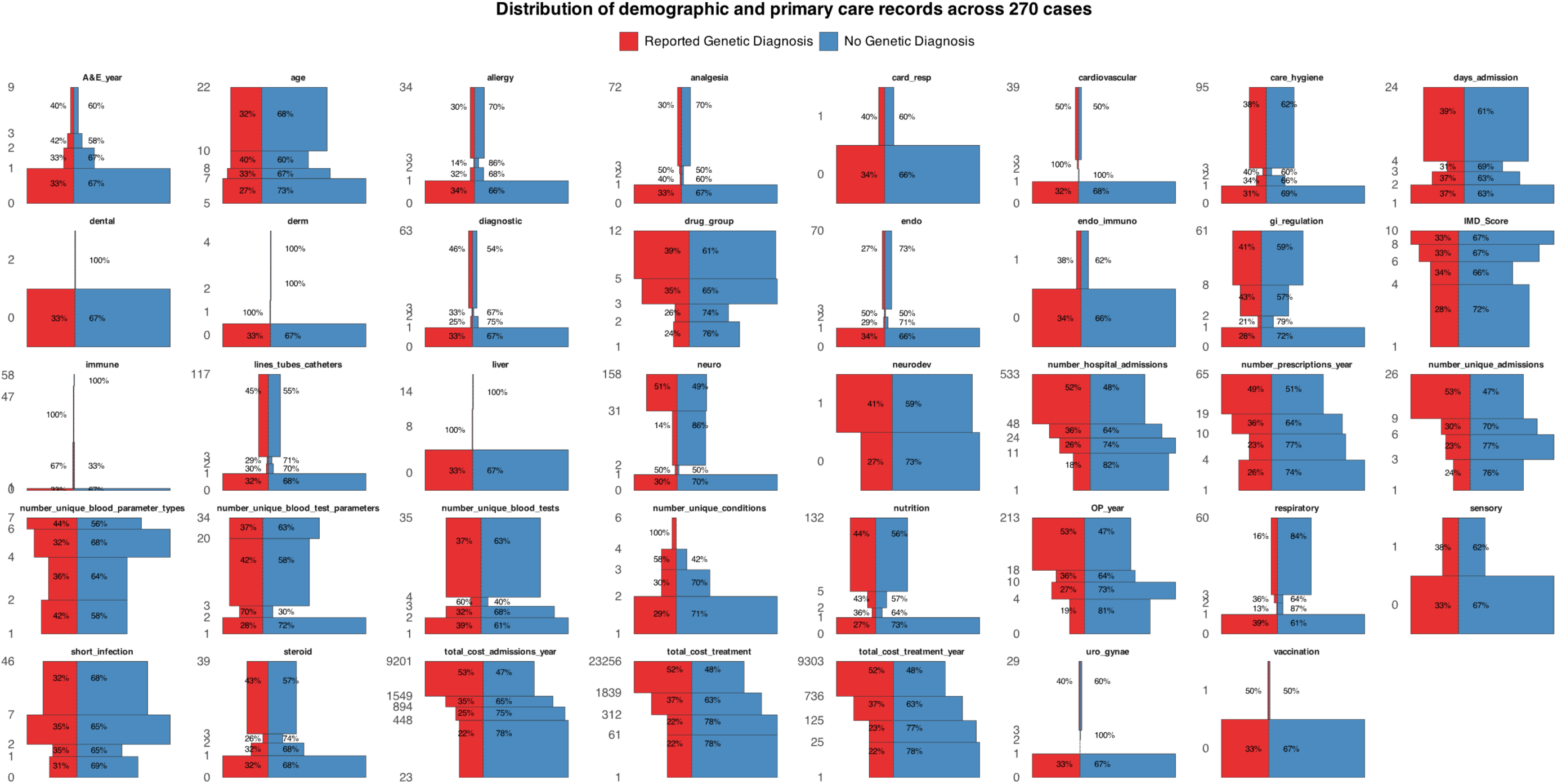

**Figure S2.**
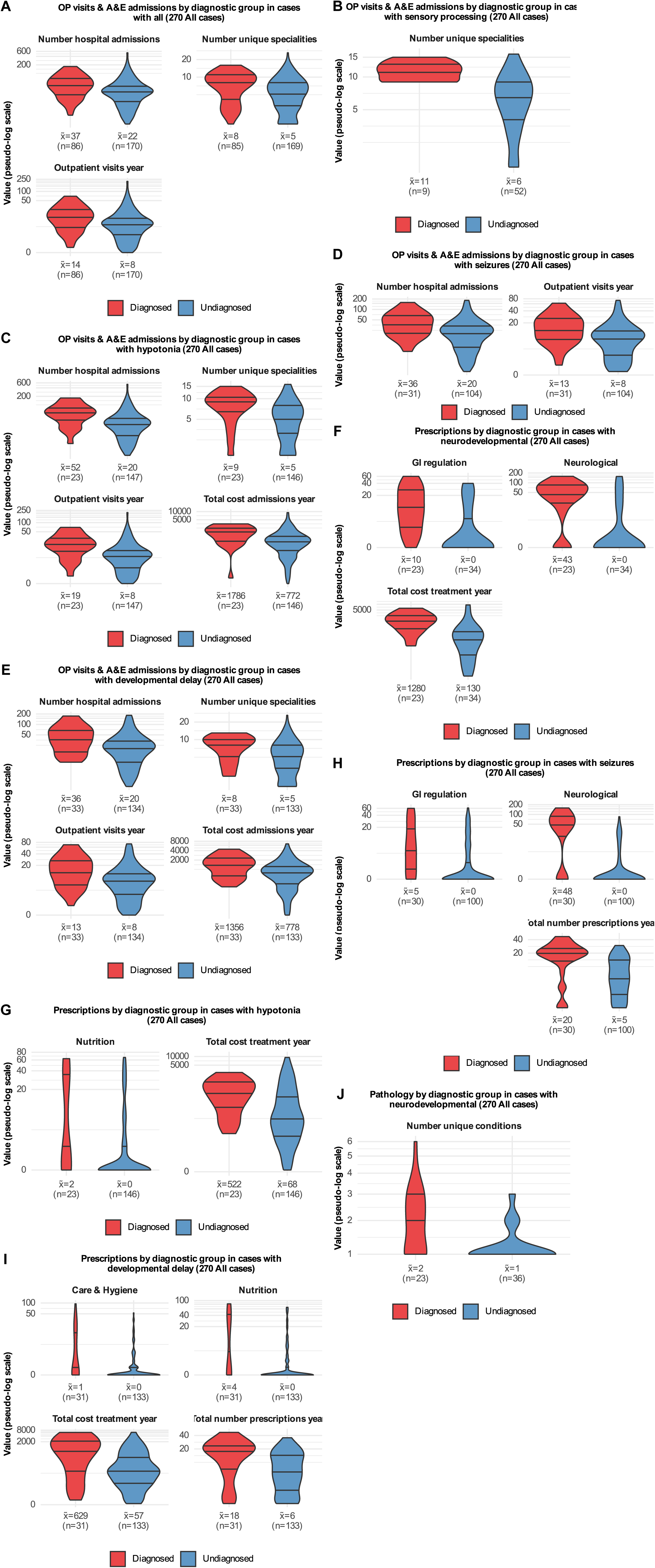

**Figure S3.**
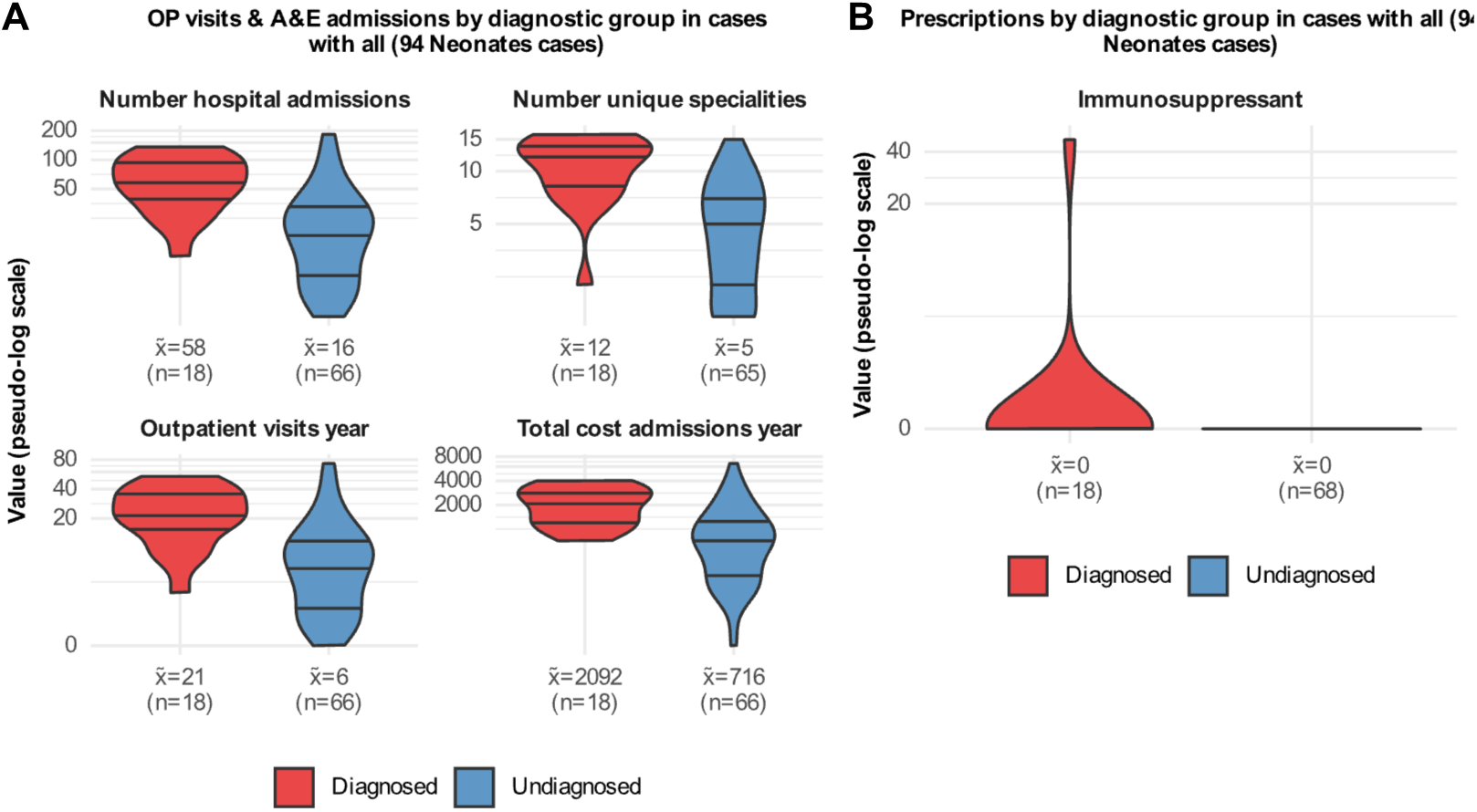

**Figure S4.**
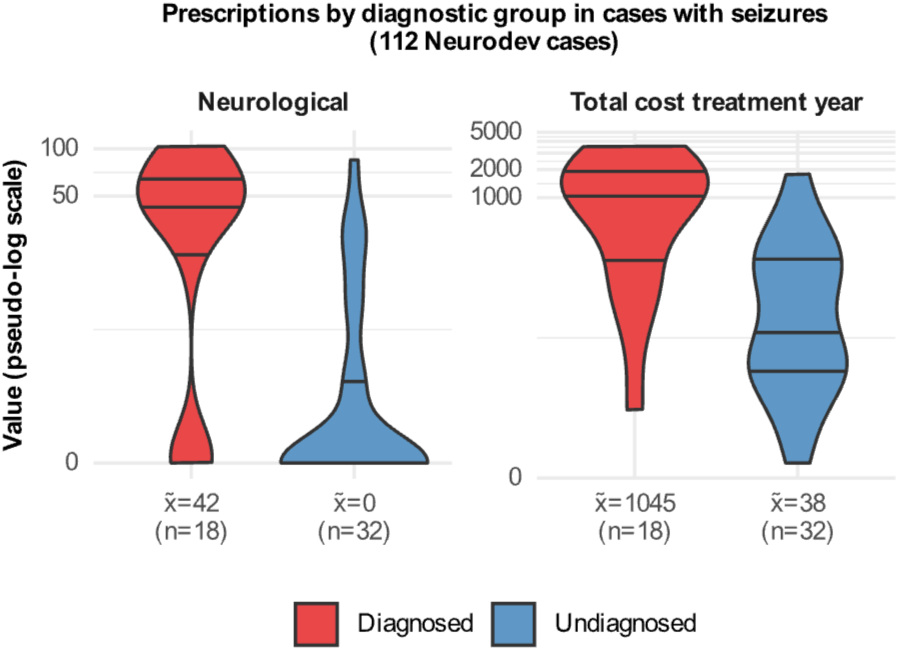

**Table S1.**

|  | Diagnosed cases |  |  |  | Undiagnosed cases |  |  |  |
| --- | --- | --- | --- | --- | --- | --- | --- | --- |
| Units | Full cohort (270) | Neonates | Neurodev | Critically III | Full cohort (270) | Neonates | Neurodev | Critically III |
| All | 87 (32 %) | 18 (19 %) | 47 (42 %) | 22 (34 %) | 183 (68 %) | 76 (81 %) | 65 (58 %) | 42 (66 %) |
| card_resp | <7 | <7 | <7 | <7 | 63 | 26 | 28 | 9 |
| endo_immune | <7 | <7 | <7 | <7 | 64 | 27 | 27 | 10 |
| neuro_dev | 23 | <7 | 12 | 9 | 36 | 18 | 11 | 7 |
| sensory | 9 | <7 | <7 | <7 | 54 | 23 | 24 | 7 |
| Hypotonia | 23 | <7 | 12 | 8 | 160 | 75 | 49 | 36 |
| Seizures | 31 | <7 | 19 | 10 | 114 | 49 | 36 | 29 |
| Developmental delay | 33 | <7 | 24 | 9 | 147 | 76 | 36 | 35 |

**Table S2.**
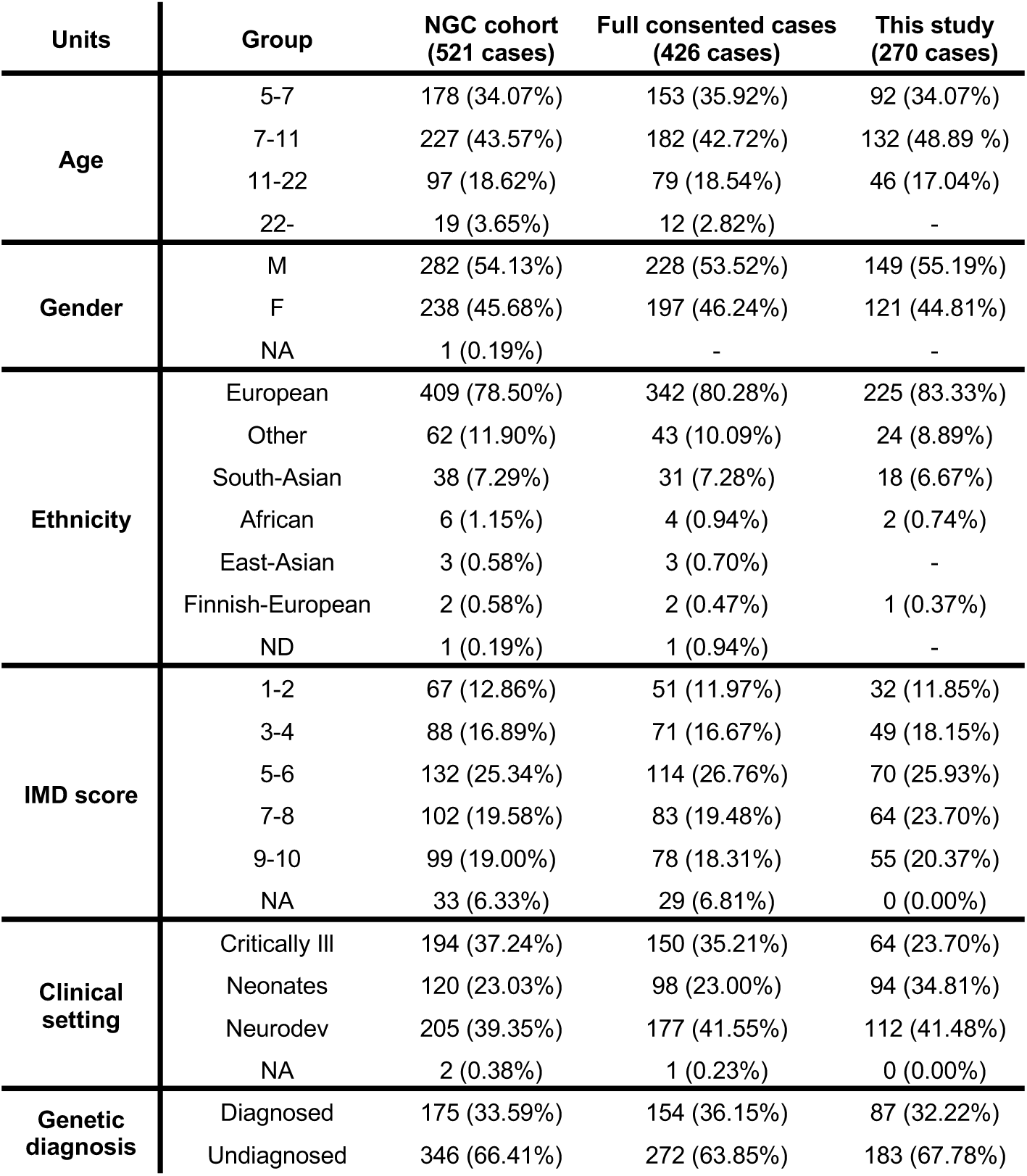

