## Supplementary material for "Long-Term Healthcare Utilization After Genomic Diagnosis in Seriously Ill Children": Suplementary_Information

**Supplementary Information**

**Supplementary Figures**

**Figure S1** Histogram of demographic and primary care features across 270 cases, stratified by diagnostic group. Each feature is discretized into four quantile-based bins to enable standardized comparisons across variables. Bars indicate the absolute number of individuals per bin, grouped by diagnostic status (diagnosed vs. undiagnosed). To enhance visibility for features with high or skewed values, the y-axis is scaled using log1p.

**Figure S2** Violin plot visualization of healthcare utilization (A-E: OP visits and A&E admissions; F-I: Prescriptions; K: Pathology), highlighting statistically significant differences ($n \geq 7$; $p < 0.001$) between diagnosed and undiagnosed groups across the cohort (270 cases) and multiple clinical conditions. Each panel corresponds to a specific condition-utilization comparison, with violin plots contrasting diagnosed (red) and undiagnosed (red) groups. The y-axis uses a pseudo-log scale to accommodate wide-ranging values. Median, interquartile range (IQR), and sample size for each group are annotated below the respective plots.

**Figure S3** Violin plot visualization of healthcare utilization (A: OP visits and A&E admissions; B: Prescriptions), highlighting statistically significant differences ($n \geq 7$; $p < 0.001$) between diagnosed and undiagnosed groups across the 94 Neonates cases and multiple clinical conditions. Each panel corresponds to a specific condition-utilization comparison, with violin plots contrasting diagnosed (red) and undiagnosed (red) groups. The y-axis uses a pseudo-log scale to accommodate wide-ranging values. Median, interquartile range (IQR), and sample size for each group are annotated below the respective plots.

**Figure S4** Violin plot visualization of healthcare utilization, highlighting statistically significant differences ($n \geq 7$; $p < 0.001$) between diagnosed and undiagnosed groups across the 112 Neurodevelopmental cases and multiple clinical conditions. Each panel corresponds to a specific condition-utilization comparison, with violin plots contrasting diagnosed (red) and undiagnosed (red) groups. The y-axis uses a pseudo-log scale to accommodate wide-ranging values. Median, interquartile range (IQR), and sample size for each group are annotated below the respective plots.

**Figure S5** Violin plot visualization of healthcare utilization, highlighting statistically significant differences ($n \geq 7$; $p < 0.001$) between diagnosed and undiagnosed groups across the 64 Critically ill cases and multiple clinical conditions. Each panel corresponds to a specific condition-utilization comparison, with violin plots contrasting diagnosed (red) and undiagnosed (red) groups. The y-axis uses a pseudo-log scale to accommodate wide-ranging values. Median, interquartile range (IQR), and sample size for each group are annotated below the respective plots.

**Supplementary Tables**

**Table S1** Distribution of cases across the full cohort and within each hospital unit, stratified by diagnostic group and the specific conditions highlighted in bold in Table 1. Shaded cells indicate small cell counts ($n<7$) subject to suppression.

**Table S2** Distribution of age, gender, ethnicity, IMD score, clinical setting and genetic diagnosis across the NGC cohort (521 cases), fully consented cases (426 cases), and the current study (270 cases).

**Phenotype-Based Genetic Diagnostic Model Pipeline**

The models were built using all available Human Phenotype Ontology (HPO) terms, demographics and primary care records (named as features) from this study. Obsolete HPO terms were manually updated using the Monarch Initiative website (https://monarchinitiative.org/). A binary classification method was developed to predict genetic diagnostic outcomes from patient phenotypic data and primary care records. This method integrates Principal Component Analysis (PCA) for data dimensionality reduction with a supervised Linear Discriminant Analysis (LDA) classification model. The complete analysis was implemented in R (v4.5.0).

Step 1: Prepare Input Data

The model requires four data inputs: a binary matrix of HPO terms, demographics and primary care records (transformed features: model input matrix) and a binary vector indicating diagnostic status (model output vector).

- *Diagnostic Status Vector*: Cases were categorized into a positive (diagnostic) group and a negative (non-diagnostic) group.
    - *Positive Group*: Included cases with 'Reported' results.
    - *Negative Group*: included cases with 'Non-diagnostic' results.
- *Binary HPO Matrix*: All HPO terms were curated using the 'ontologyIndex' R package to ensure consistency, with obsolete terms updated to their current valid equivalents. The curated set of terms was retained for downstream analysis. HPO terms more specific than level 3 in the ontology's hierarchy were replaced by their corresponding ancestor at level 3. Terms at or broader than level 3 were retained. This process resulted in a binary matrix representing the presence or absence of normalized HPO terms for each case (rows) and HPO term (columns).
- *Demographics*: Sex and ethnicity were converted to binary vectors. Age was divided into three quantile-based intervals using breakpoints at 6 and 11 years-old and IMD scores were divided into five quantile-based intervals using breakpoints at the 2, 4, 6 and 8. This transformation allowed to convert these features into binary matrices.
- *Primary care*: Numeric features were divided into four quantile-based intervals using breakpoints at the 0th, 25th, 50th, 75th, and 100th percentiles, rounded up for interpretability. Features with four or fewer unique values were one-hot encoded. Missing values were encoded as zeros. This transformation allowed to convert these features into binary matrices.

Step 2: Optimize Number of Principal Components (PCs) via bootstrapping

To find the optimal number of PCs for the model, a bootstrapping procedure is performed by iterating over a candidate range of PC counts. For each candidate PC count, 100 bootstrapped iterations are run as follows:

- *Sample Data*: 70% of the data is sampled for training, ensuring a balanced class distribution is maintained.
- *Apply PCA*: PCA is performed on the binary feature matrix of the training sample using the 'prcomp' function, without centering or scaling the data.
- *Train LDA Model*: An LDA model is trained on the PCA-transformed training data. The model is built to generate a classification probability vector classifying cases into their predefined

diagnostic groups using the 'lda' function from the 'MASS' package (v2.2.2) with default parameters.

*OR*

- *Predict and Evaluate:* The complete dataset is projected into the PCA space derived from the training sample. The trained LDA model is then used to predict the genetic diagnostic status for the full dataset.
- *Store Results:* The Area Under the Curve (AUC) for the iteration is calculated from the Receiver Operating Characteristic (ROC) curve using the 'roc' function from the 'pROC' package (v1.18.5). The AUC and the PCA + LDA model from the iteration are stored.

Step 3: Select Optimal Number of PCs
The best PC count is identified by calculating the average AUC across the 100 iterations for each tested PC count. The number of PCs that yields the highest average AUC is selected as the optimal number.

Step 4: Select Final Model
From the 100 models trained using the optimal number of PCs (determined in Step 3), the single model whose AUC is closest to the median AUC of that set is selected as the final model.

Step 5: Determine Best Probability Threshold
Using the predictions from the final model selected in Step 4, the optimal probability threshold is identified. This threshold is the point that maximizes the sum of Sensitivity and Specificity.

Step 6: Final Output
This pipeline generates the final, optimized components for the PCA + LDA genetic diagnostic model:

- Optimal number of PCs
- Selected PCA+LDA model
- LDA scores
- Optimal classification probability threshold

**Feature Contribution Analysis Pipeline**

The contribution of input features to the model's predictions was assessed to determine the influence of individual clinical features. This protocol describes the specific methodology for calculating this contribution.

Step 1: Define Input Data
The following data and a pre-trained model are required for the analysis:

- *Normalised HPO terms, demographic data and primary care records matrix*: A binary matrix, HPO_terms, where rows represent individual cases and columns represent each of the input features. The values within the matrix indicate the presence or absence of a specific feature for each case.
- *Trained PCA + LDA model*: The combined PCA and LDA model trained for classification.

Step 2: Initialize Change Matrix

This step creates a matrix to store the results of the analysis.

- A change_matrix is generated with dimensions corresponding to the input data.
- All values in this matrix are initialized to 0. It will store the calculated difference in prediction probabilities.

Step 3: Iterative Contribution Calculation

This step systematically calculates the change in the model's prediction probability by removing one feature at a time for each case.

- *Outer Loop*: For each feature j:
- *Inner Loop*: For each case i:
  - Check for term presence: If feature[i, j] == 1, proceed with the following steps. Otherwise, skip to the next case.
  - *Get original classification probability*:
    - Apply the PCA + LDA model to the original feature vector for case i (feature[i, :]).
    - The resulting prediction probability is stored as the original probability (oProb_matrix[i, j]).
  - *Create modified case*:
    - Create a temporary copy of the case's feature vector, feature_modified[i, :].
    - In this copy, set the value for term j to 0 (feature_modified[i, j] = 0).
  - *Get Modified Classification Probability*:
    - Apply the PCA + LDA model to the feature_modified [i, :] vector.
    - The resulting prediction probability is stored as the modified probability (mProb_matrix[i, j]).
  - *Calculate classification probability change*:
    - Store the difference between the original and modified probabilities in change_matrix[i, j] = oProb_matrix[i, j] - mProb_matrix[i, j].

Step 4: Generate Final Output

The result of this pipeline is an aggregated score for each feature, representing its average influence on the model's output.

- For each feature j, calculate the average contribution by taking the mean of all values in change_matrix[:, j] where the term was originally present (feature[i, j] == 1).
- Calculate the respective 95% confidence intervals for each average contribution score.

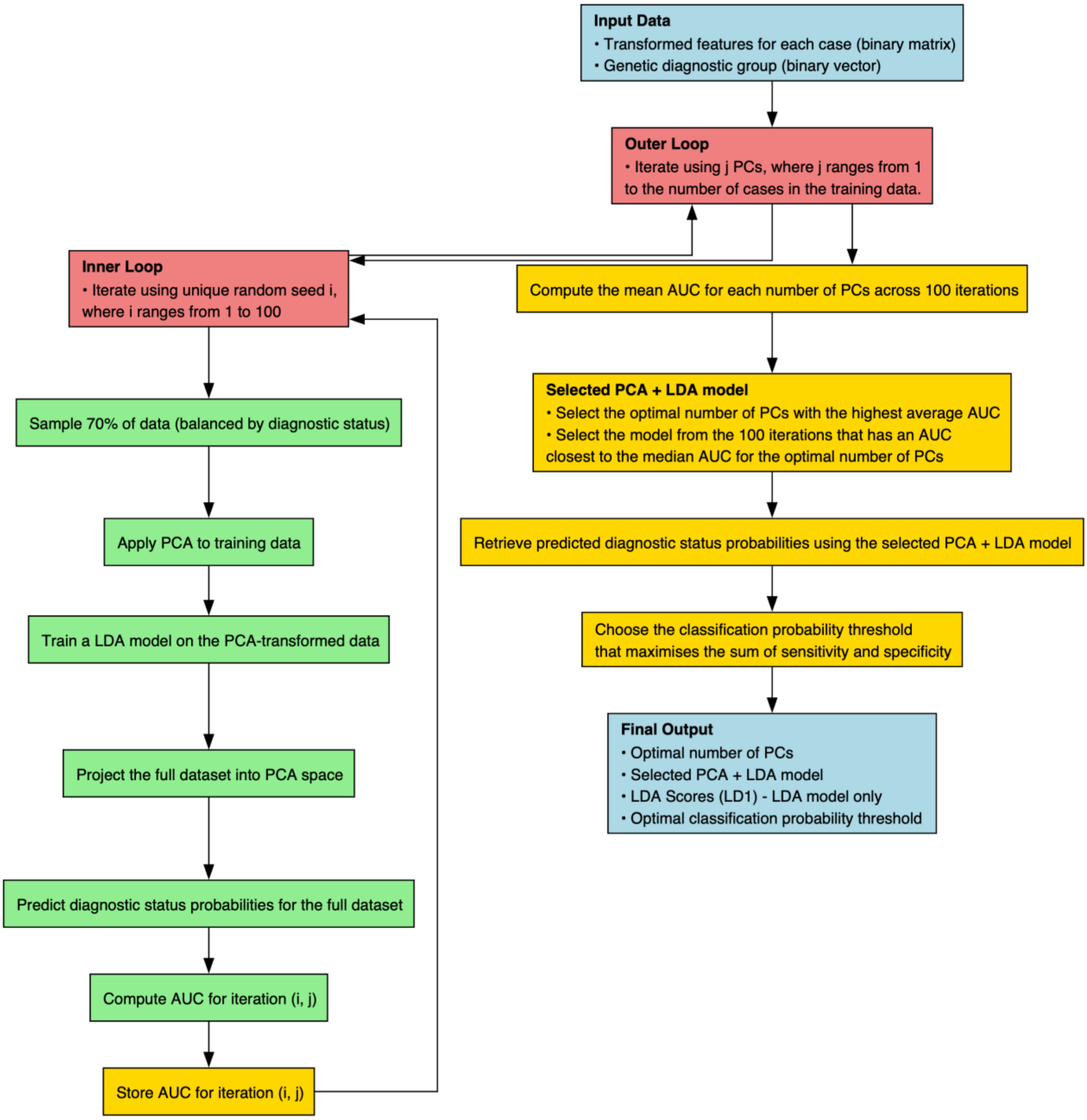


This diagram illustrates the process of predicting diagnostic status from the transformed features, using PCA for dimensionality reduction and LDA for classification. The methodology employs a nested loop structure to determine the optimal number of principal components and select a final, robust model. Each step is visually distinguished by colour: blue boxes represent the initial input data and final outputs; red boxes indicate the outer loop (iterating through the number of PCs) and the inner loop (100 bootstrapped iterations); green boxes show the core computational tasks; and yellow boxes mark the supporting tasks for result aggregation and model selection.

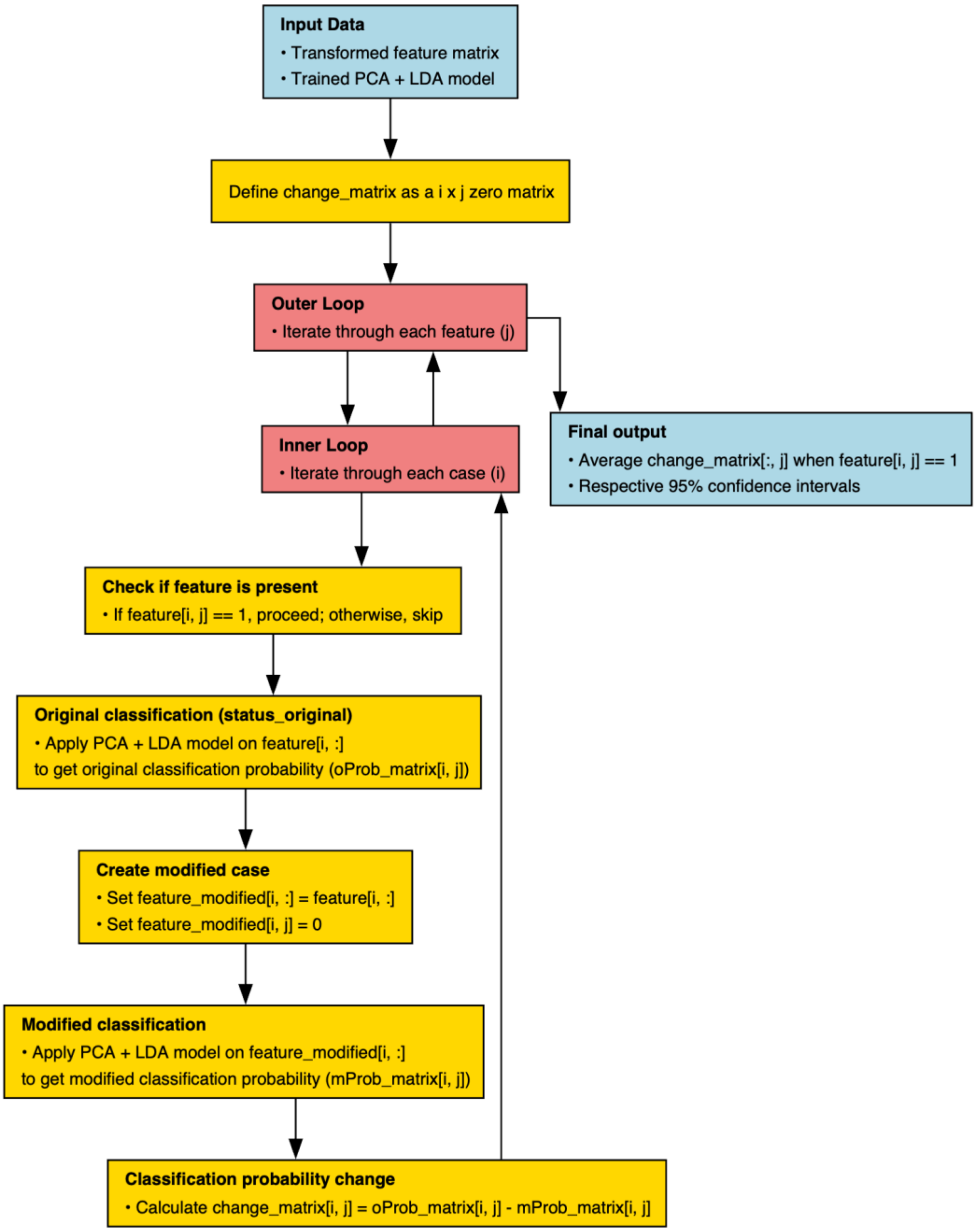


This flowchart details the process of quantifying the impact of removing each transformed feature on the classification probability generated by the selected PCA + LDA model. The methodology uses a nested loop to iterate through each term and each case. For cases where a term is present, the change in classification probability upon its removal is calculated. The final output is the average of these changes for each term, providing a measure of its overall diagnostic importance. Each step is visually distinguished by colour: blue boxes represent input and output, red boxes indicate loops, and yellow boxes show the core computations and supporting tasks.
